# Longitudinal modeling of multimorbidity trajectories using large language models

**DOI:** 10.1101/2024.10.02.24314786

**Authors:** Lu Yang, Elliot Bolton, Gowri Nayar, Russ B. Altman

## Abstract

Multimorbidity, the co-occurrence of two or more chronic conditions within an individual, is a major and escalating global health challenge, complicating treatment regimens, straining healthcare resources, and worsening patient outcomes. The complex interplay of shared genetic predispositions, biological pathways, and socioeconomic factors underpins its development, but clinical and research efforts have largely focused on managing diseases in isolation. Understanding multimorbidity trajectories—the accumulation and interaction of chronic diseases over time—is essential to improving preventive strategies and optimizing personalized care. Here, we introduce ForeSITE (Forecasting Susceptibility to Illness with Transformer Embeddings), a novel, transformer-based framework that harnesses advanced machine learning to predict multimorbidity progression. By analyzing longitudinal data from 480,000 participants in the UK Biobank, ForeSITE identifies distinct patterns in the co-occurrence and timing of diseases. Our temporal disease network provides insights into how certain diseases might share common genetic, environmental, or socioeconomic factors, offering more specific guidance for earlier detection and more effective disease management.

## Introduction

Precision medicine, with its focus on tailoring treatments to individual patients, has revolutionized modern healthcare by integrating diverse health data sources. These data include biobanks like the UK Biobank, which offer comprehensive longitudinal datasets, as well as electronic health records (EHRs), genomics, proteomics, metabolomics, and lifestyle data. This vast and diverse data enables insights into disease progression and the underlying biological mechanisms. For example, the UK Biobank’s detailed genetic and clinical data facilitates associations between genetic variants and complex conditions, such as asthma and Alzheimer’s disease, while considering modifiable risk factors like diet and physical activity. However, while precision medicine has enabled targeted therapies for individual diseases, a major challenge remains in predicting and managing multimorbidity—the development of multiple chronic conditions over a patient’s lifetime. Current approaches tend to treat diseases in isolation, missing critical insights into how conditions develop together. To address this, our research leverages advanced machine learning models, specifically transformer-based language models^1^, to predict multimorbidity trajectories, offering a more comprehensive understanding of disease interactions and progression over time. Using data from 480,000 participants in the UK Biobank, we aim to forecast disease progression across the full spectrum of health outcomes. For example, through patient stratification, we can identify individuals who experienced childhood asthma and are at increased risk of developing eczema within 5 years, allergic rhinitis in 10 years, and later adult conditions like osteoporosis or anxiety disorders within 20 years. By pinpointing these high-risk patients early, clinicians could intervene sooner, potentially delaying disease onset or slowing progression. This kind of stratification offers a clear path toward more proactive and personalized care, making treatment not only more effective but also more efficient. Ultimately, this approach holds the potential to transform standard care practices by aligning interventions with patients’ individual health profiles and risks.

Multimorbidity, characterized by the simultaneous presence of two or more chronic conditions within an individual, is an escalating global challenge that poses complex questions about how diseases interact and evolve over time^2^. Beyond merely identifying which diseases tend to co-occur, a deeper understanding of when these conditions develop in relation to one another is critical. Temporal analysis focuses on the specific timing and sequence of disease onset, providing insights into how the occurrence of one condition might influence the development of another. Longitudinal analysis, on the other hand, examines how these diseases progress and interact over the long term within individuals, allowing researchers to track health changes across extended periods. Our research leverages large language models (LLMs) to study and predict disease trajectories, combining both temporal and longitudinal aspects. By using data from biobanks like the UK Biobank, LLMs enable us to model the timing and progression of multiple conditions, offering a more comprehensive understanding of how multimorbidity evolves. This dual approach helps inform how healthcare interventions can be better timed to prevent or mitigate the impact of multimorbidity. Addressing these challenges is particularly critical for socioeconomically disadvantaged populations, who often experience earlier onset of multimorbidity and face worse outcomes, such as reduced mobility, diminished quality of life, and increased mortality^3,4^. By leveraging these insights, we aim to guide the development of more effective healthcare strategies that account for both the complexity and timing of disease development in multimorbidity.

The challenge posed by multimorbidity is particularly pronounced in the elderly population, where it emerges as a pervasive and complex dilemma in healthcare. Managing multiple chronic conditions simultaneously introduces challenges such as balancing conflicting treatments, addressing medication interactions, and coordinating care across different specialists^5^. Although often conflated with “comorbidity”, these terms describe distinct phenomena. Comorbidity, introduced by Feinstein in 1970, refers to the presence of a primary disease alongside one or more secondary conditions^6^. This focus on a main condition can appeal to specialists treating specific diseases but often results in a segmented approach within healthcare systems^7^. As a result, care becomes fragmented, which is particularly harmful for patients managing multiple chronic illnesses^7^. The interaction between multiple coexisting conditions often complicates treatment decisions, underscoring the need for a more holistic and integrated approach^8,9^. By prioritizing the broader concept of multimorbidity, healthcare providers can examine the interconnections between all concurrent conditions, rather than addressing each in isolation. In this work, we focus on multimorbidity as a central area of inquiry. This integrated perspective offers a comprehensive view of individual health outcomes, without prioritizing any single disease over others, thereby better capturing the complexity of real-world patient experiences.

Investigating the longitudinal trajectory and patterns of multimorbidity in health and well-being is crucial, especially given the aging global population^10^. By analyzing the prevalence, progression, and interplay of various medical conditions, researchers can identify key risk factors and develop more effective strategies for preventing and treating diseases^11,12^. Multimorbidity facilitates the identification of these risk factors by uncovering patterns of disease co-occurrence, tracing how multiple conditions evolve over time, and highlighting shared genetic, lifestyle, or environmental factors that contribute to the development of multiple diseases across the lifespan. Numerous studies have examined trends and characteristics of multimorbidity, but most have been limited to analyzing a relatively small set (fewer than 100) of diseases, such as cardiovascular diseases, myocardial infarction, and depression. Common methods, such as factor analysis and cluster analysis, are often employed to investigate multimorbidity patterns^13–18^, while other approaches aim to summarize the non-random correlations among individual diseases^19–21^. However, many of these studies tend to overlook critical temporal details, focusing primarily on directionality without accounting for the timing between key clinical events such as birth, disease onset, or mortality. Therefore, it is essential to evaluate the intervals between illnesses within populations and to understand how different health conditions are related over time.

In this work, we introduce ForeSITE, a framework based on a GPT-style model, to predict diagnostic events for 479,769 individuals in the UK Biobank, which includes time-stamped data of first diagnoses from hospital inpatient records. This comprehensive dataset offers a unique opportunity to study multimorbidity by examining the temporal sequence of disease onset across a large population. Using this framework, we present a methodology to extract temporal relationships among 1,129 disease entities. Our algorithm demonstrates superior performance compared to state-of-the-art language models in predicting the onset of a comprehensive range of phenotypes. Additionally, we construct a disease temporal network based on associations derived from ForeSITE and compare it with a static human disease network from the UK Biobank. ForeSITE offers a general-purpose approach for diagnostic event prediction and can be applied to other biobanks that include dated health-related data.

## Methods

### Overview of multimorbidity trajectories modeling

We extracted individual diagnostic events from the UK Biobank and processed them into sequences of ICD-10 codes based on the time of their first occurrences. This dataset allowed us to develop **ForeSITE**, a GPT-style model designed to predict individuals’ subsequent diagnostic events based on their patient histories. To conduct an in-depth analysis of multimorbidity trajectories, we constructed a disease network in which the edges represent the temporal duration between one disease and the next. The overall strategy of our work is illustrated in Figure 1. This network provides invaluable insights into the evolving patterns and relationships among various diseases, contributing to a more comprehensive view of the dynamics of multimorbidity.

**Figure 1:**
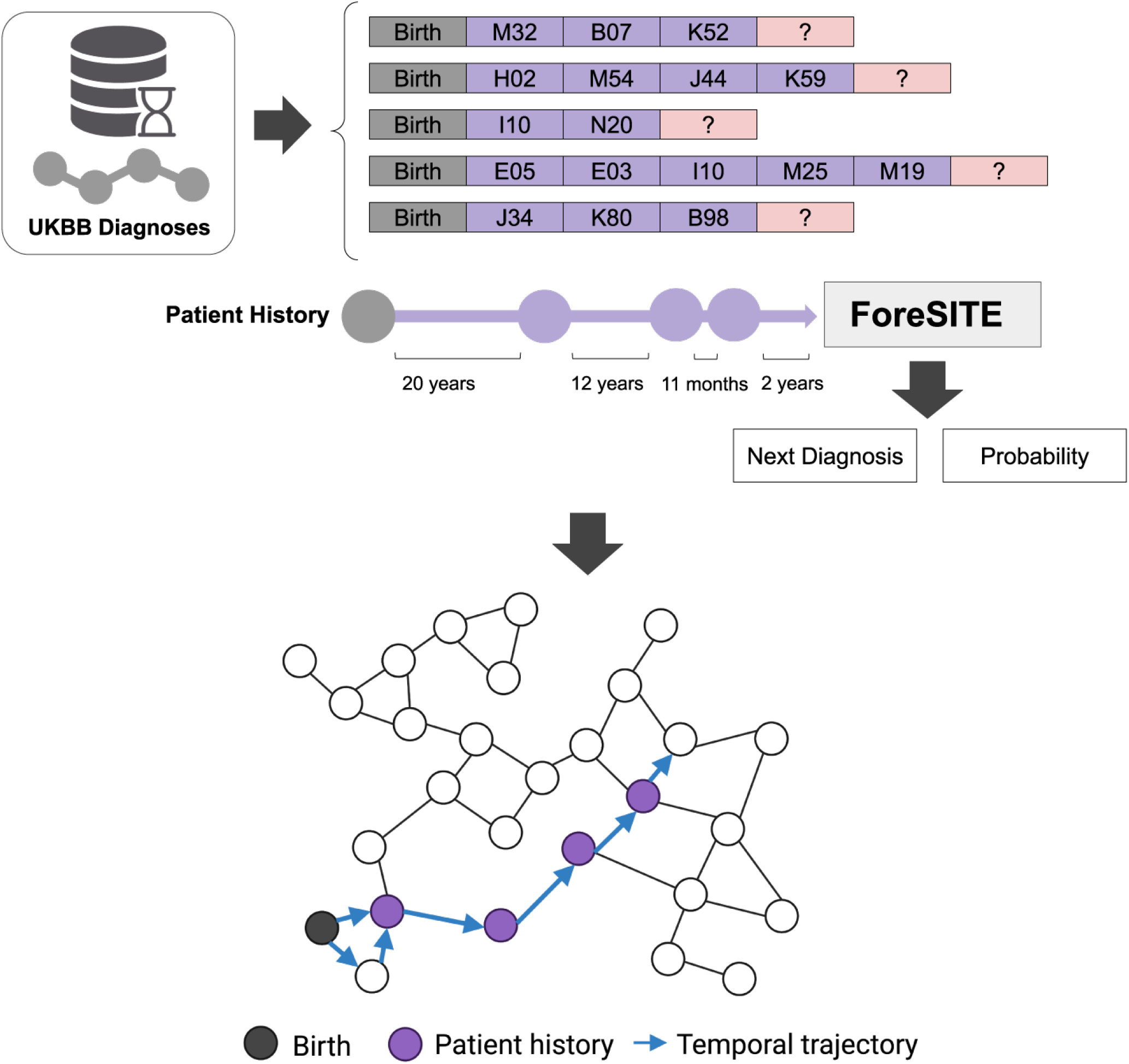
Overview of modeling multimorbidity trajectories. A series of patient diagnostic sequences was extracted from the UK Biobank, encompassing both birth and diagnostic events, with corresponding time durations between diagnoses highlighted in violet in the upper plot. A subsequent network analysis of these clinical events provides insight into the temporal trajectories of diseases, illustrating the evolving patterns and interconnections of various medical conditions over time.

### Sequential diagnoses data cohort

The UK Biobank (UKBB) is a substantial longitudinal dataset, containing health information for approximately half a million individuals in the United Kingdom, providing a robust foundation for studies into disease pathogenesis. We extracted specific health-related outcomes from this dataset, marked by the “first occurrence” of any 3-character ICD-10 code. This process yielded diagnostic codes and their corresponding first recording dates for each individual, sourced from various records such as primary care, hospital inpatient data, death registers, and self-reported medical conditions.

Out of 481,151 diagnostic sequences, we retained 479,769 individuals with valid, time-stamped first occurrences of diseases and corresponding birth dates. Figure 2 visually represents an example of these dated diagnostic records. To illustrate the temporal progression between disease onset, we crafted nine unique time codes (Table 1) and converted the historical data of 1,129 disease entities into sequential diagnostic codes paired with corresponding time intervals. The UKBB anonymized and encrypted the data before release for analysis. We then partitioned this sequential diagnoses data into training (80%), validation (10%), and testing (10%) sets, ensuring no overlap between patients across the training and validation sets. The testing set was reserved solely for evaluating the performance across different models. In our prediction framework, we developed the model to forecast both the timing of the next diagnostic event and the ensuing phenotype code, providing insights into the complexities of disease development and progression.

**Table 1:** Time codes designed for UK Biobank sequential diagnoses data.

| Parameter | Setting |
| --- | --- |
| T1 | <2 weeks |
| T2 | 2 weeks – 1 month |
| T3 | 1 – 3 months |
| T4 | 3 – 6 months |
| T5 | 6 months – 1 year |
| T6 | 1 – 2 years |
| T7 | 2 – 5 years |
| T8 | 5 – 10 years |
| T9 | >10 years |

**Figure 2:**
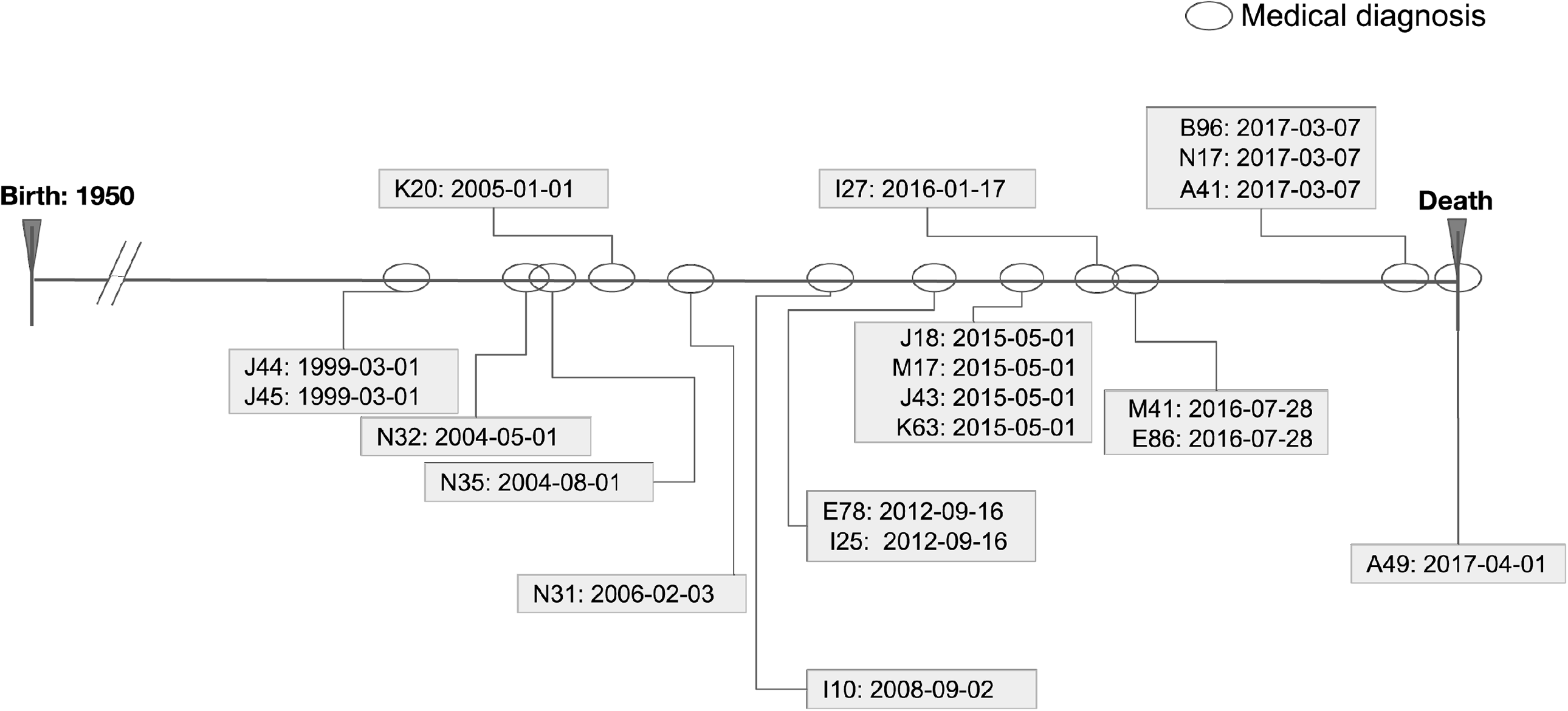
Schematic of dated health-related outcomes in UK Biobank. This illustration depicts the diagnostic timeline for a hypothetical patient, highlighting key medical diagnoses. Each diagnosis is represented by a circle, recorded at the time of first occurrence, providing a coherent visual record of the patient’s medical history.

### ForeSITE framework

For diagnostic event prediction, we utilized a GPT-style model fine-tuned on **BioMedLM**^22^, as shown in Figure 3. ForeSITE is an autoregressive, decoder-only Transformer model with a customized vocabulary that accommodates all 3-character ICD-10 codes. The specific configurations and adjustments applied to this model are detailed in Table 2. This tailored approach leverages contemporary language processing techniques to enhance the model’s precision in predicting diagnostic sequences in clinical settings.

**Table 2:** GPT-2 model architecture settings.

| Parameter | Setting |
| --- | --- |
| Hidden Size | 768 |
| Layers | 12 |
| Attention Heads | 12 |
| Vocab Size | 30,044 |
| Sequence Length | 1024 |

**Figure 3:**
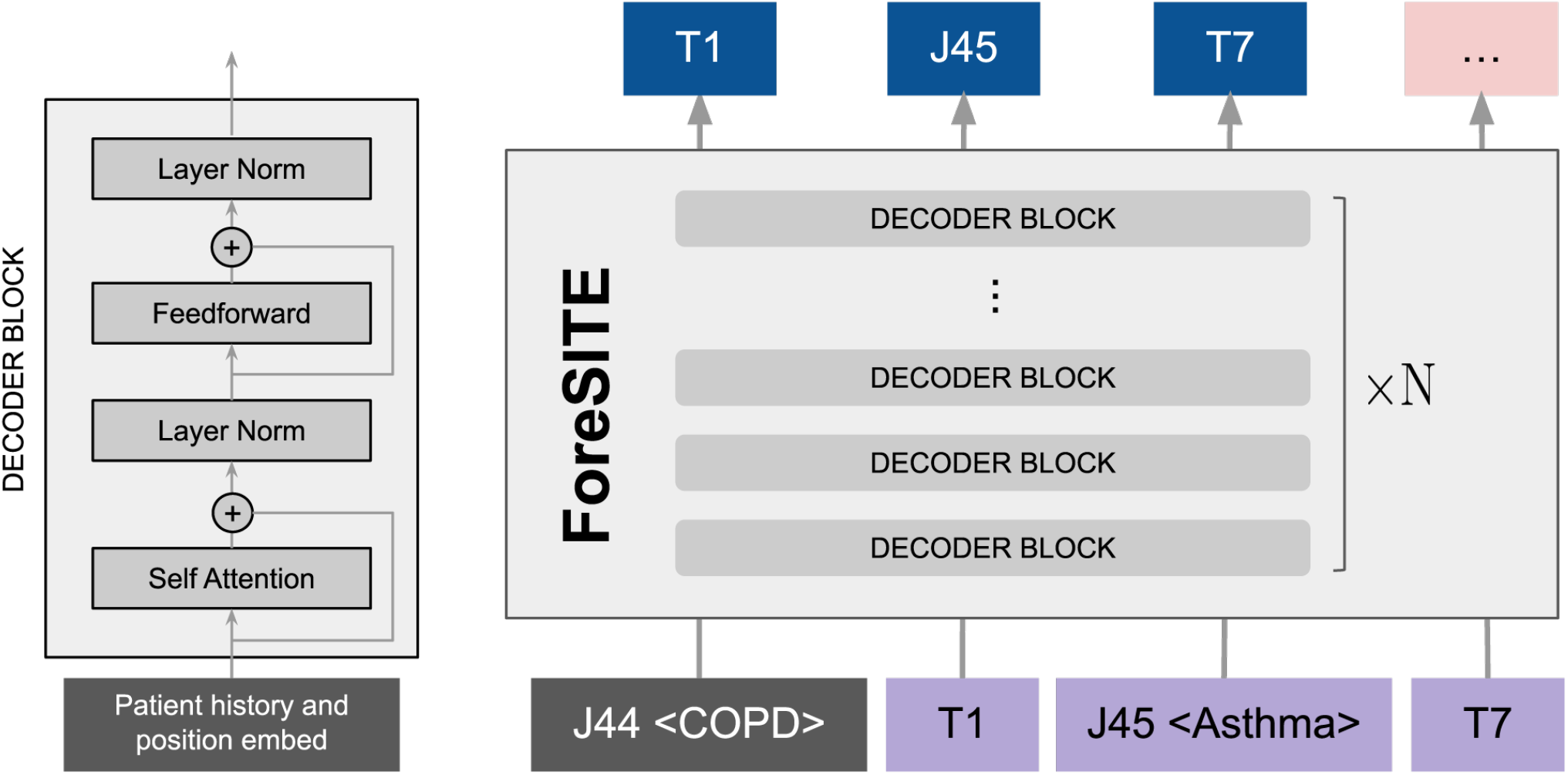
ForeSITE framework overview. A GPT-style architecture adapted for analyzing patient diagnostic sequences. The model was fine-tuned using BioMedLM^22^, incorporating detailed descriptions for each phenotype code and corresponding 3-character ICD-10 codes. This approach enhances the model’s accuracy in interpreting complex medical data.

### Phenotype prediction

As illustrated in Figure 3, each occurrence of a time code or phenotype code in an individual’s diagnostic timeline triggers a prediction. A prediction is considered correct if it exactly matches the corresponding ground truth event. To achieve this, we employed four distinct sequence models: a long short-term memory (LSTM) network^23,24^, a recurrent neural network (RNN)^25^, a GPT-style model^26,27^, and the fine-tuned BioMedLM^22^. These models were trained using PyTorch in Python^28^ for computational efficiency. For evaluation, we performed a comparative analysis using top-*k* precision metrics to assess the models’ ability to predict diagnostic timelines accurately.

### Network construction

Our analysis employs both a static network constructed using WGCNA (Weighted Gene Co-expression Network Analysis) methodology^29,30^ and a temporal trajectory network. In the constructed disease network, individual nodes represent specific diseases, and an edge between two nodes is created if the likelihood of these diseases co-occurring differs significantly from what would be expected if they occurred independently.

We quantify the association between two diseases using the phi coefficient, calculated as shown in Equation 1:

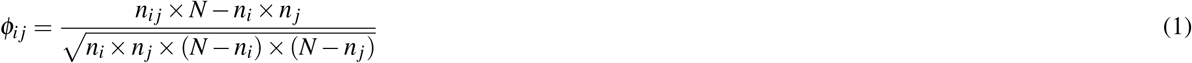

where *φ*_*ij*_ is the phi coefficient measuring the association between diseases *i* and *j*; *n*_*ij*_ represents the number of individuals diagnosed with both diseases *i* and *j*; *n*_*i*_ and *n* _*j*_ are the numbers of individuals diagnosed with diseases *i* and *j*, respectively; and *N* is the total number of individuals in the dataset.

For the temporal network analysis, we incorporated event probabilities as determined by our ForeSITE framework instead of using static correlations in comorbidity. To determine connectivity, we introduce a threshold *τ*, derived from the scale-free topology criterion^21,29,30^. Two phenotypes are deemed to have a strong association if connected by an edge, as formalized in Equation 2:

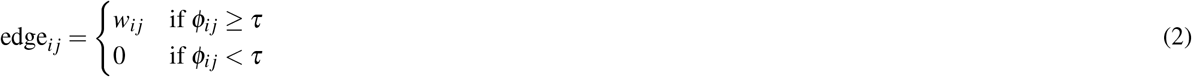

where edge_*i j*_ is the weight of the edge between diseases *i* and *j*, and *w*_*ij*_ is the calculated weight or strength of association between diseases *i* and *j* (e.g., set equal to *φ*_*ij*_ or derived from it).

## Results

### Sequential diagnoses data analysis

A total of 479,769 unique individuals were selected for our study, as detailed in Table 3. Our analysis of the UK Biobank data illustrates a clear association between multimorbidity and age (Figure 4A). Specifically, the number of diagnosed disorders per individual increases with age. Notably, while no patients at the age of 10 have more than five diagnostic codes, over 90% of those aged 80 have been diagnosed with more than five diseases. The distribution of diagnostic sequence lengths within the UK Biobank (Figure 5) reveals an average sequence length of 24.8 events (including time codes), with a maximum length of 251. The age of onset varies considerably across different diseases. Figure 4B presents histograms of ages at first occurrence for individuals with six specific conditions, illustrating that certain diseases, such as asthma, can manifest at any age, whereas others, like dementia, are primarily diagnosed after age 50.

**Table 3:** Summary characteristics for training, validation, and testing sets.

|  | Training Set<br>(N=383,825) | Validation Set<br>(N=47,943) | Testing Set<br>(N=47,938) |
| --- | --- | --- | --- |
| Sex (%) |  |  |  |
| Female | 207,451 (54.0) | 26,299 (54.8) | 26,113 (54.4) |
| Male | 176,374 (46.0) | 21,644 (45.2) | 21,825 (45.6) |
| Age (%) |  |  |  |
| ≥ 65 | 294,051 (77.0) | 36,968 (77.1) | 37,096 (77.4) |
| < 65 | 89,774 (23.0) | 10,975 (22.9) | 10,842 (22.6) |
| Ethnicity (%) |  |  |  |
| White | 357,925 (93.3) | 45,099 (94.0) | 45,110 (94.0) |
| Mixed | 2,173 (0.6) | 298 (0.6) | 303 (0.6) |
| Asian | 6,169 (1.6) | 733 (1.5) | 769 (1.6) |
| Black | 7,240 (1.9) | 962 (2.0) | 926 (1.9) |
| Chinese | 1,129 (0.3) | 143 (0.3) | 140 (0.3) |
| Other | 3,365 (0.9) | 428 (0.9) | 430 (0.9) |
| Unknown | 1,258 (0.3) | 169 (0.4) | 144 (0.3) |
| Did not answer | 789 (0.2) | 111 (0.2) | 116 (0.2) |

**Figure 4:**
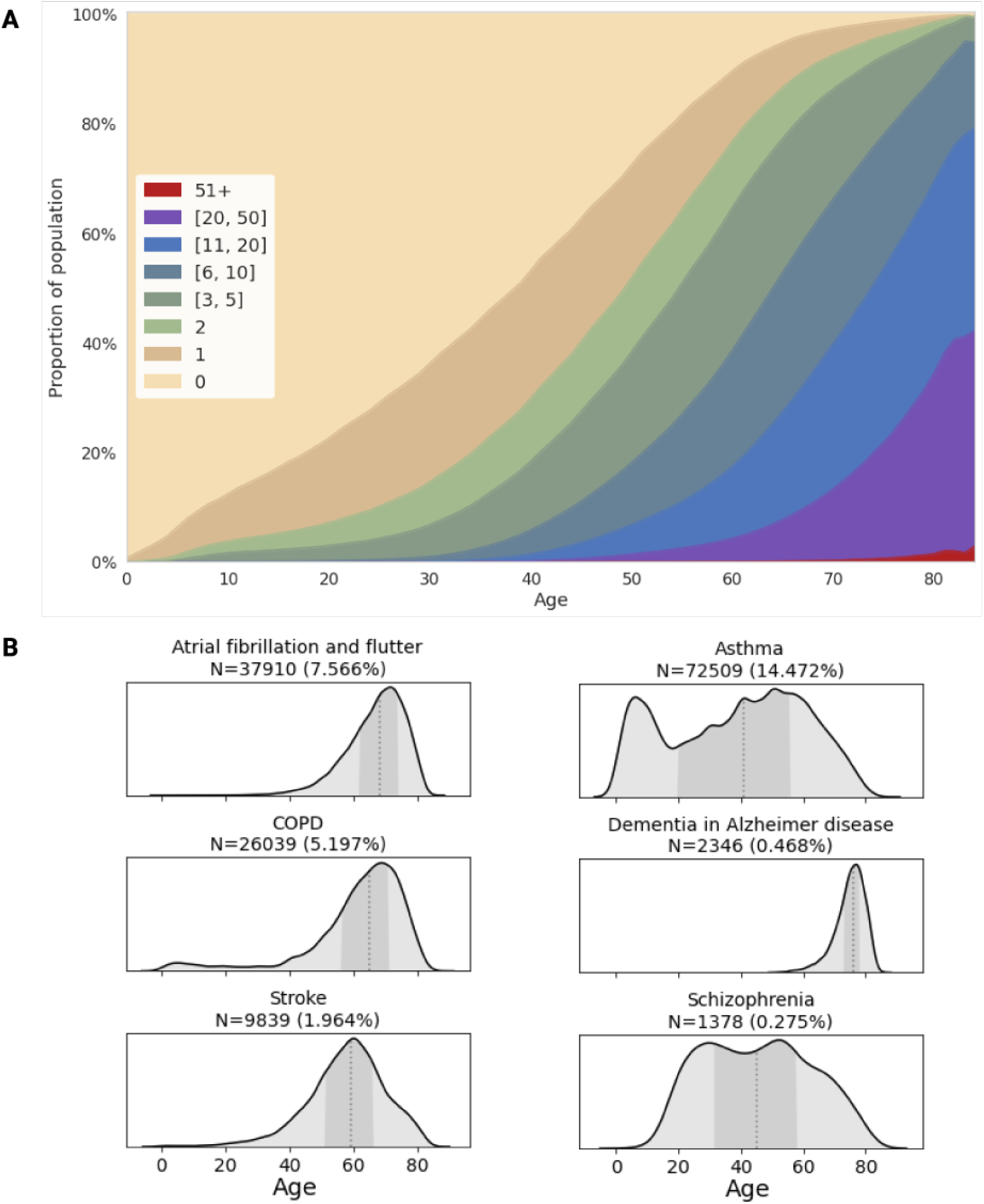
An overview of multimorbidity in the UK Biobank population. (A) Proportion of individuals with multimorbidity across different age groups, showing a progressive increase with advancing age. (B) Histograms illustrating the age of first occurrence for six distinct disorders: (1) atrial fibrillation and flutter, (2) asthma, (3) chronic obstructive pulmonary disease (COPD), (4) dementia in Alzheimer’s disease, (5) stroke, and (6) schizophrenia.

**Figure 5:**
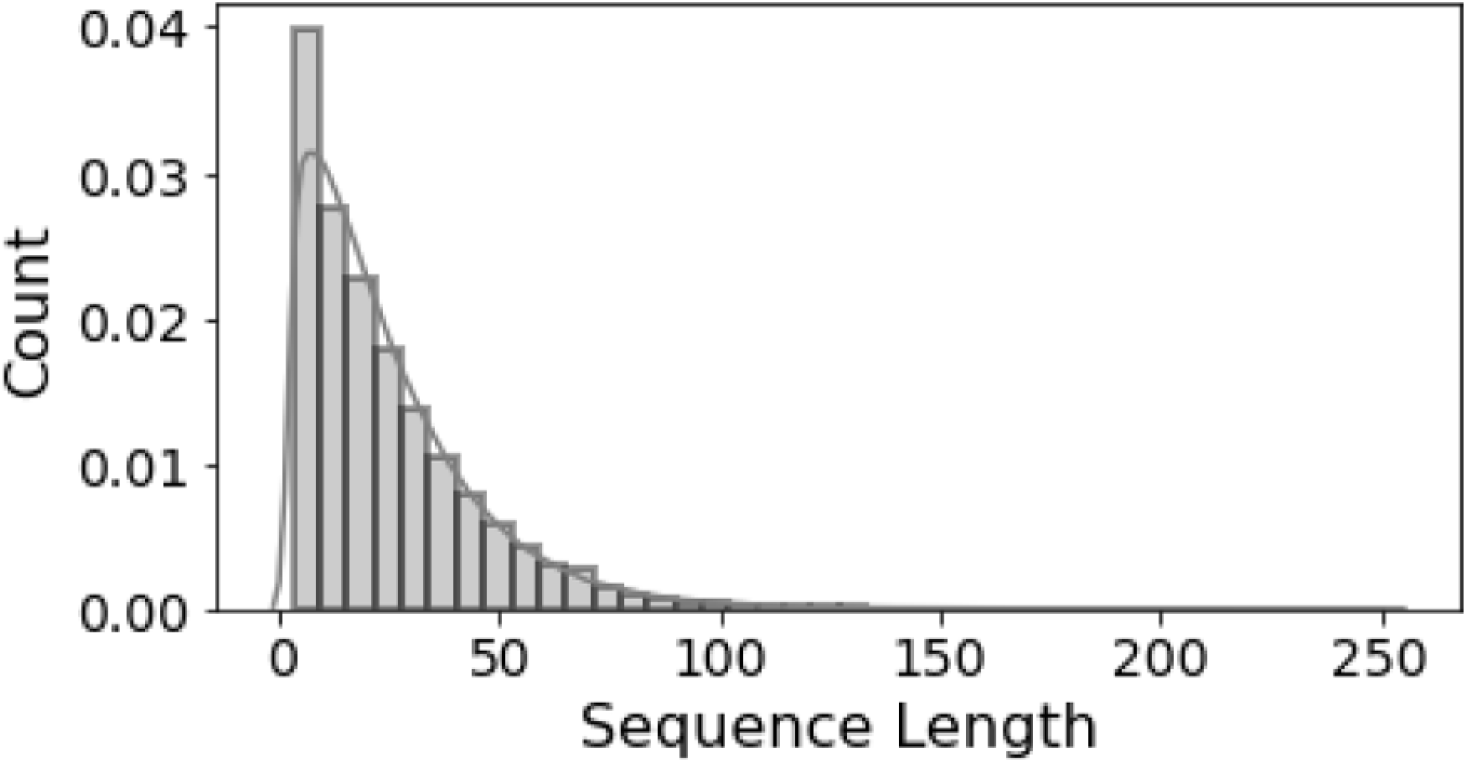
Distribution of diagnostic sequence lengths in the UK Biobank. This histogram illustrates the distribution of event codes among individuals in the UK Biobank. The *x*-axis represents the number of event codes assigned to each person, reflecting the complexity of health-related occurrences. The *y*-axis denotes the number of individuals possessing a specific count of event codes, revealing how frequently certain sequence lengths occur within the population.

We divided our dataset into training (383,825 patients), validation (47,943 patients), and testing (47,938 patients) sets. Analysis of key demographics—sex, age, and ethnicity—revealed that the subsets share similar characteristics. The overall study population comprises approximately 54% women, with a predominance of elderly patients aged 65 and over (77%), and individuals identifying as white (94%) (Table 3).

### ForeSITE Performance

We predicted both time codes and diagnostic events following birth for individuals in the UK Biobank dataset. Among the various models tested—including RNN, LSTM, GPT, and our ForeSITE model—the GPT-2 style architecture adapted from BioMedLM demonstrated the highest predictive performance (Figure 6). Specifically, ForeSITE achieved top-1, top-10, top-20, top-30, and top-50 accuracy scores of 25.9%, 68.18%, 74.21%, 78.21%, and 83.60%, respectively. Here, “top-*k* accuracy” refers to the proportion of instances where the correct diagnosis was among the model’s top *k* predictions. Since multiple subsequent diagnostic events can follow any given event, the top-1 accuracy provides insight into the most likely next disease.

**Figure 6:**
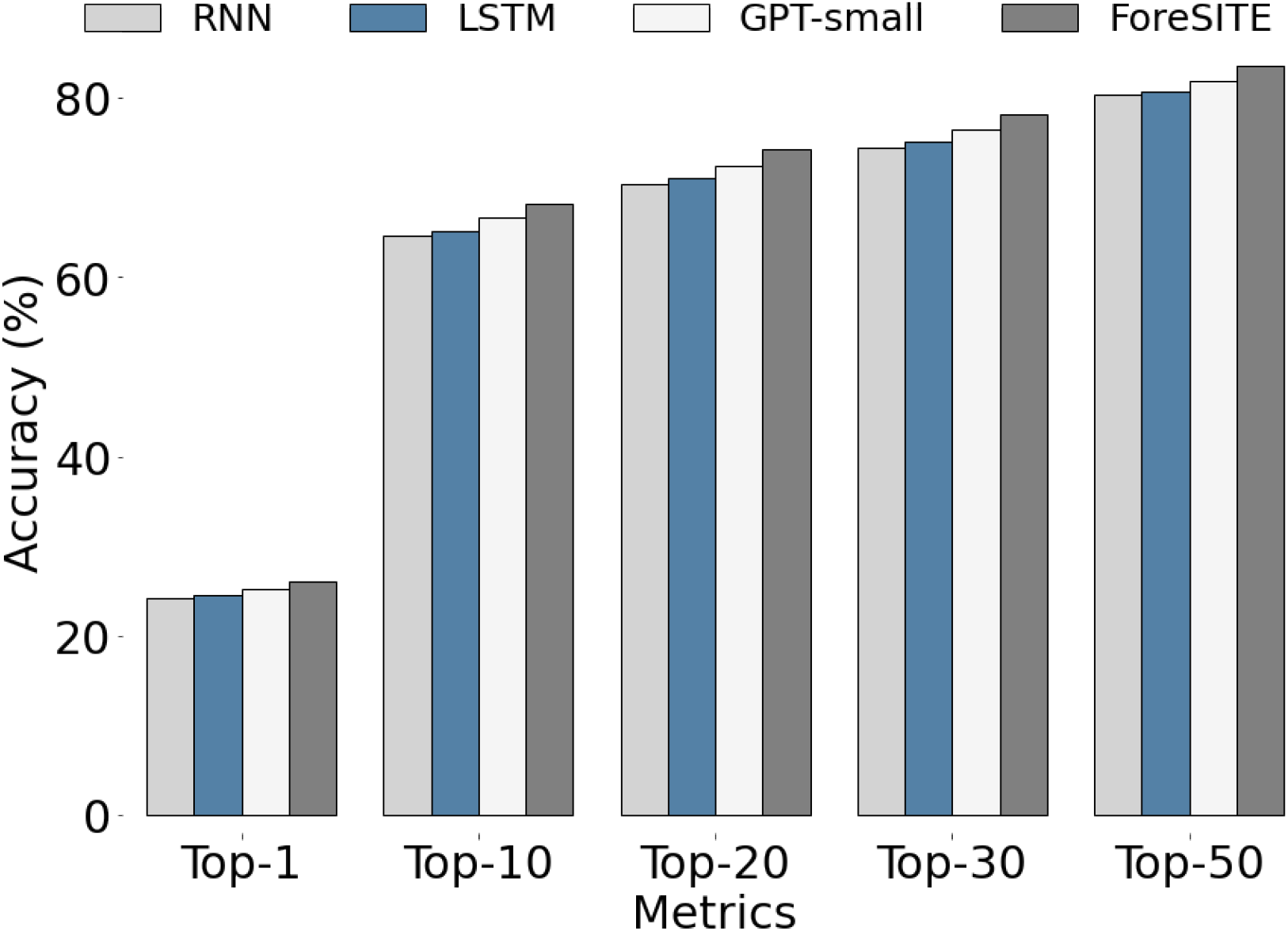
ForeSITE prediction for future diagnostic events. Comparison of top-*k* performance among different language models (RNN, LSTM, GPT, and ForeSITE) in predicting future diagnostic events in the UK Biobank. The ForeSITE model demonstrates superior performance across all top-*k* metrics.

### Human disease co-occurrence network

We constructed a comorbidity network to represent disease co-occurrences within the UK Biobank (Figure 7). This network consists of 301 disease nodes connected by 426 edges, offering insights into the complex relationships among different medical conditions. Certain disease codes exhibit high connectivity; for example, hypertension (I10) is connected to multiple other diseases, indicating its common co-occurrence with various conditions. The average node degree of 1.42 suggests a moderate level of interconnection among diseases. Common diseases are highlighted in Supplementary Figure S2, and the line weights in the network indicate the strength of association between diagnoses—for instance, asthma (J45) and COPD (J44) often co-occur.

**Figure 7:**
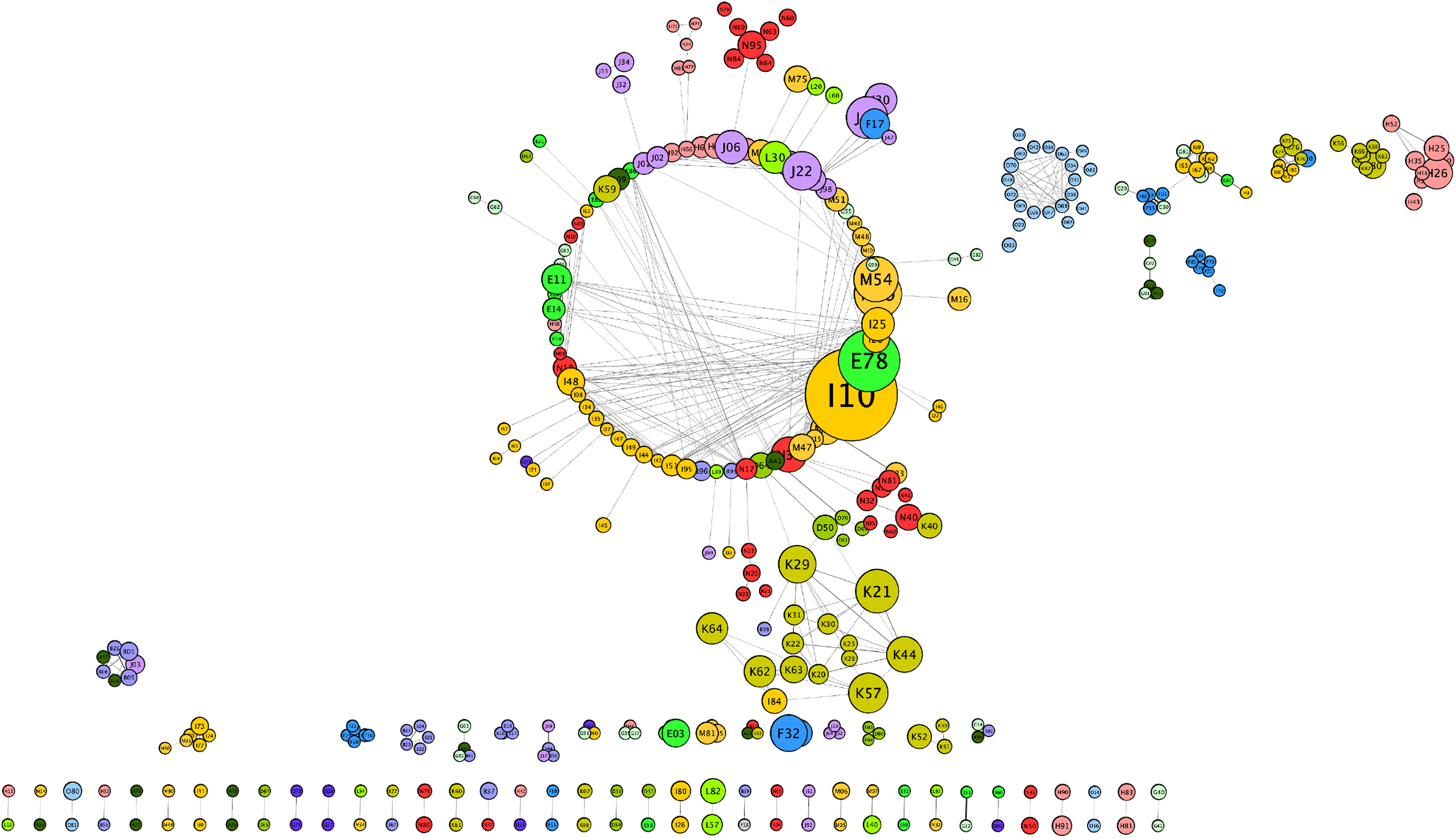
Comorbidity network constructed based on disease co-occurrences in the UK Biobank. In this network, node size is determined by the number of patients corresponding to each disease. Each node is labeled with the relevant 3-character ICD-10 code and colored to represent its specific disease category. Edges between two nodes are formed if the probabilities of the two diseases co-occurring deviate from what would be expected if they were independent. This structure visually captures the complex interplay of diseases within the population.

Suppelmentary Figure S1 demonstrates that the co-occurrence network follows a scale-free topology, where a few nodes have many connections while most have few. While this comorbidity network helps identify patterns of disease co-occurrence, it does not provide insights into the temporal progression from one disorder to another.

### Temporal human disease trajectory network

Using ForeSITE predictions, we constructed a directed temporal disease network that elucidates the complex relationships among phenotypes over time. This network captures not only which diseases are related but also the typical time intervals between their occurrences. By differentiating diseases closely related in time from those more distant, we gain a richer understanding of disease progression patterns beyond simple pairwise relationships. The network is heterogeneous, comprising directed disease networks across various time constraints, detailed in Supplementary Figures S3, S4, S5, S6, S7, S8, S9, S10.

A segment of the multimorbidity trajectory network, displaying the most likely disease trajectories for a selection of chronic conditions, is presented in Figure 8. This graph provides an overview of connections between diseases such as chronic obstructive pulmonary disease (COPD, J44), asthma (J45), emphysema (J43), kidney failure (N17), neuromuscular dysfunction of the bladder (N31), hypertension (I10), osteoarthritis of the knee (M17), and sepsis (A41). For example, hypertension typically emerges more than 15 years post-birth, with disorders associated with lipoprotein metabolism (E78) commonly appearing 1-2 years before hypertension onset. The directed temporal structure of the network uncovers detailed multimorbidity patterns, offering crucial insights into the timing and progression of chronic diseases and their co-occurrence.

**Figure 8:**
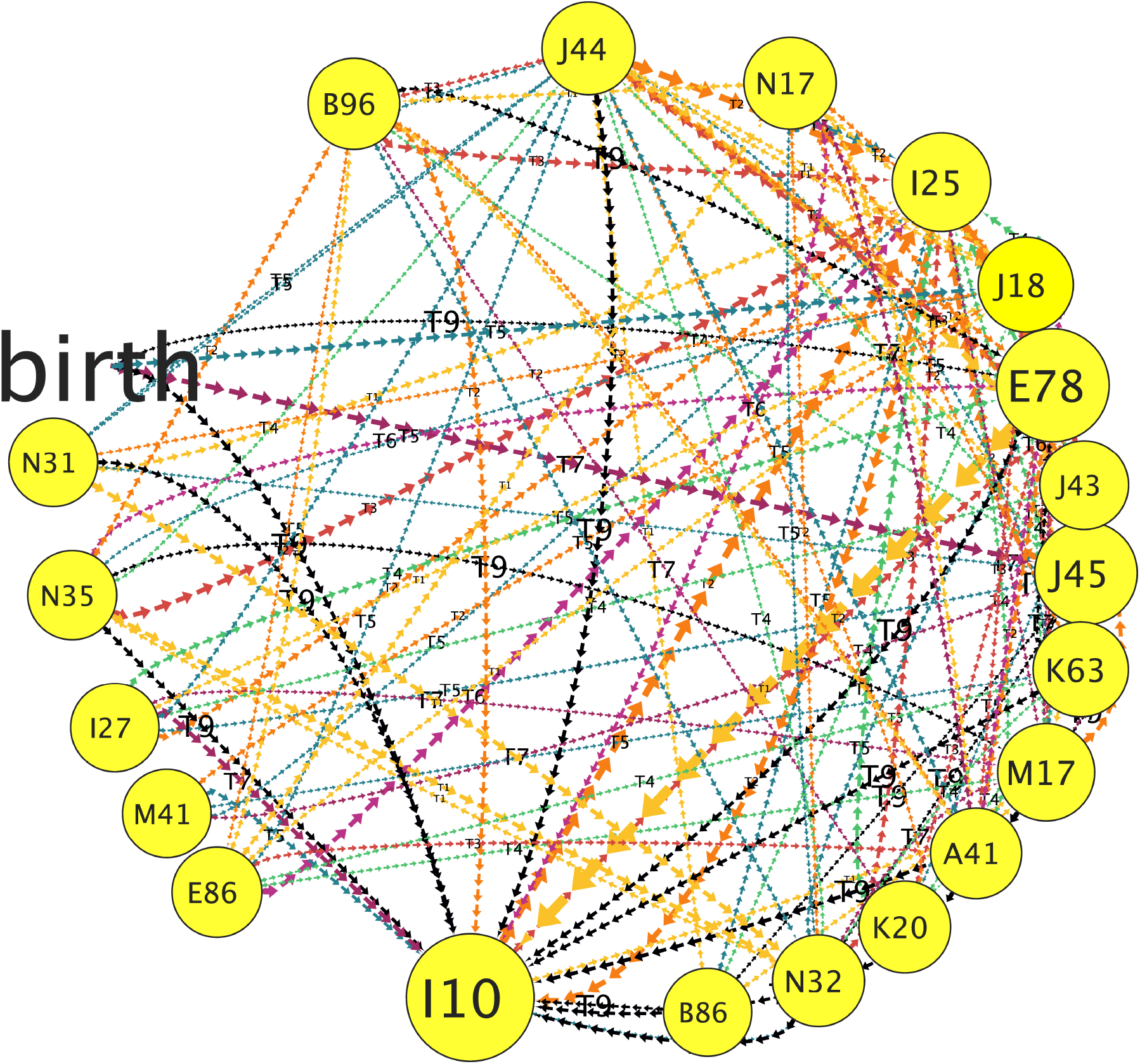
Temporal network of disease trajectories. Visualization of the interconnected trajectories of selected chronic diseases over time, based on ForeSITE predictions. Nodes represent diseases, and directed edges indicate the progression from one disease to another within specific time intervals.

### Web user interface for synthetic disease trajectories

We have developed a user-friendly web interface for ForeSITE to enhance interaction and demonstrate the model’s potential applications. Through this platform, users can generate synthetic disease trajectories based on our GPT-style model’s predictions. Figure 9 illustrates this feature, showcasing the generation of a synthetic patient trajectory where a diagnosis of whooping cough (A37) occurs 1-2 years post-birth.

**Figure 9:**
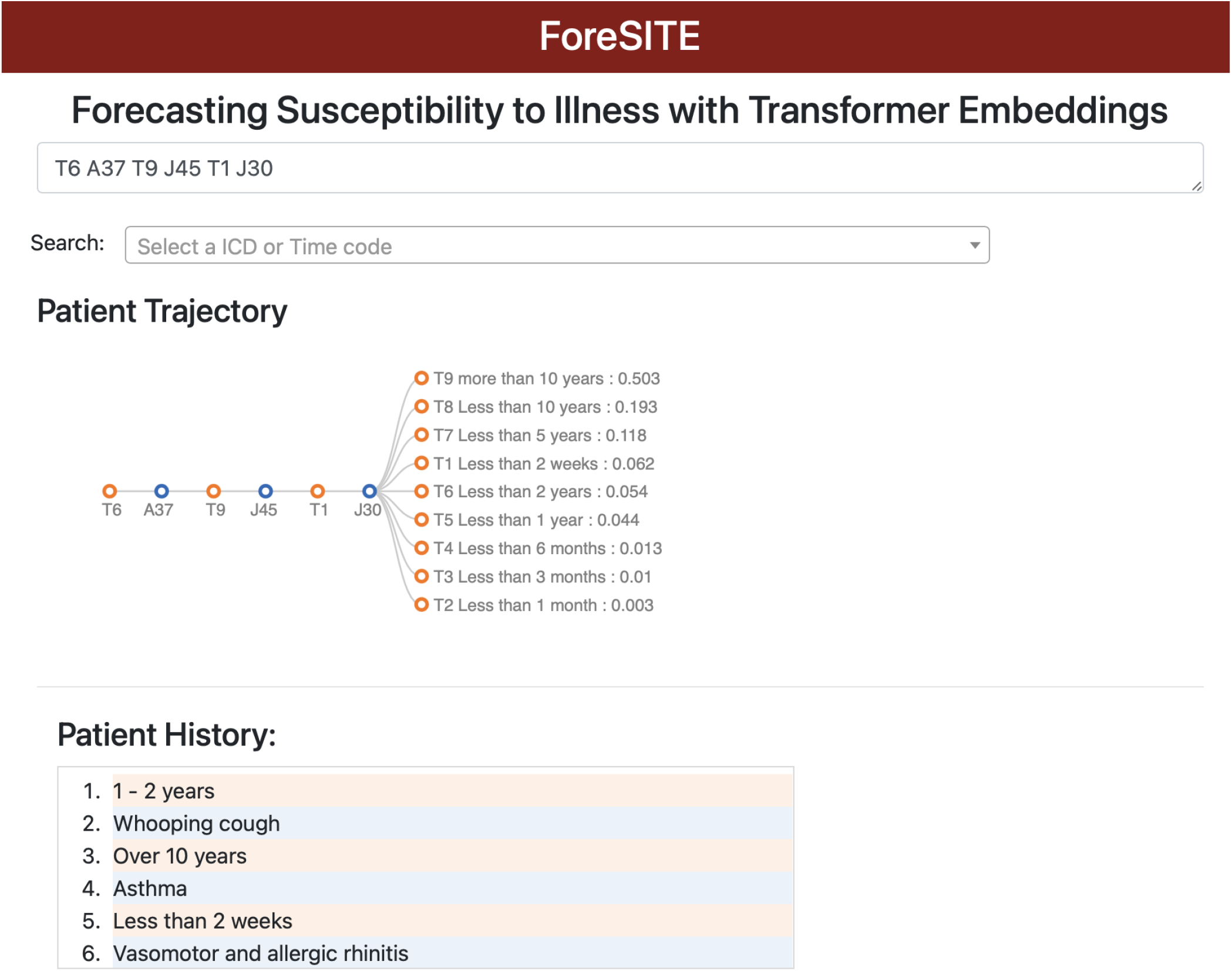
User interface for ForeSITE. A screenshot of the web interface allowing users to generate synthetic disease trajectories based on ForeSITE predictions. In this example, a synthetic patient trajectory is generated with a diagnosis of whooping cough (A37) appearing 1–2 years post-birth.

## Discussion

As the global population ages, the increasing prevalence of multimorbidity poses a significant challenge for healthcare systems, underscoring the need for effective management strategies for at-risk individuals^2^. Patients with multiple chronic conditions, referred to as multimorbid, face a higher risk of hospitalization, which contributes substantially to healthcare costs^31,32^.

Unfortunately, existing guidelines and most disease management programs are designed to address individual diseases in isolation, leaving multimorbidity largely unaddressed^33^. Furthermore, research on preventing multimorbidity remains limited. Our work in predictive modeling of disease trajectories, based on sequential diagnoses in the UK Biobank, offers a promising avenue for identifying patients who may benefit from earlier care interventions.

In this study, we performed an analysis of multimorbidity within the UK Biobank dataset, with a particular focus on age as a critical factor (Figure 4). Our findings reveal a rapid increase in the number of chronic conditions per individual with advancing age. For instance, 40% of individuals over 80 have been diagnosed with more than 20 different conditions in the UK Biobank. Leveraging the comprehensive data available, we explored the longitudinal patterns of multimorbidity, recognizing that current definitions and methodologies for analyzing disease co-occurrence often lack temporal precision. By developing clinically relevant time codes, we extracted temporal disease trajectories using a GPT-style framework, enabling more accurate predictions of individual diagnostic events. Additionally, we constructed and compared two distinct human disease networks—static and temporal—highlighting the nuanced progression of multimorbidity. These analyses provided new insights into the sequence of disease onset and progression, potentially revealing shared genetic, environmental, or lifestyle risk factors. These findings open avenues for novel therapeutic and preventive interventions.

Despite the advances made, this research is not without limitations. One of the primary challenges lies in the need to better integrate genetic risk factors into the disease trajectory modeling. While we successfully identified temporal patterns and co-occurrences of diseases, we did not fully incorporate critical determinants such as individual genetic predisposition, socioeconomic status, and lifestyle factors. Future research should aim to include these variables to offer a more comprehensive understanding of the complex progression of multimorbidity. Another limitation arises from the nature of the data itself, specifically the reliance on Electronic Health Records (EHRs) from the UK Biobank. These records are dependent on the documentation practices of healthcare professionals, which may vary in completeness and consistency across regions. Consequently, the quality of the EHR data could introduce biases or gaps in the understanding of disease trajectories. Addressing these limitations in future studies will be essential for improving the robustness and applicability of our findings, paving the way for more personalized and effective healthcare interventions.

ForeSITE represents a tool that transcends the limitations of focusing on individual diseases. By leveraging the predictive power of a GPT-style model, it allows for precise forecasting of future phenotypes across diverse population groups. This enables the exploration of complex temporal relationships between diseases, uncovering intricate connections that might otherwise remain hidden. Beyond identifying these associations, the predicted relationships form the basis for constructing a predictive disease network, designed to anticipate the development of multimorbidity. Such a network can guide preventative strategies and targeted interventions, adding a proactive dimension to patient care. However, while the results of ForeSITE are promising, they should not be viewed as a comprehensive solution. Important factors such as genetic interactions, environmental exposures, and lifestyle choices were not fully integrated into the current model. The challenge moving forward lies in incorporating these diverse elements to achieve a more holistic understanding of disease progression. By addressing this challenge, **ForeSITE** has the potential to drive the future of personalized healthcare, adapting to the unique complexities of each patient’s health journey.

## Data Availability

All data produced in the present study are available upon reasonable request to the authors.

https://www.ukbiobank.ac.uk/enable-your-research/apply-for-access

## Acknowledgements

This work was supported by NIH GM102365 and the Chan-Zuckerberg Biohub. We would like to thank the Stanford NLP group and The Center for Research on Foundation Models (CRFM), with special thanks to Prof. Percy Liang, for providing the computing resources that supported this research.

## Author contributions statement

Lu Yang conceived the original idea, extracted the data, led the experiments, and conducted all analyses. Jason E. Boltman assisted in training the ForeSITE model. Gowri Nayar contributed to the web user interface. Russ B. Altman supervised the project.

## Additional information

The methods are released under the Stanford University academic license. For commercial use or other licensing inquiries, please contact Stanford’s Office of Technology Licensing.

## Notes

### Competing Interest Statement

The authors have declared no competing interest.

### Author Declarations

Ethics committee/IRB of UK Biobank gave ethical approval for this work

