## Supplementary for "Longitudinal modeling of multimorbidity trajectories using large language models"

### 1 Supplementary Figures

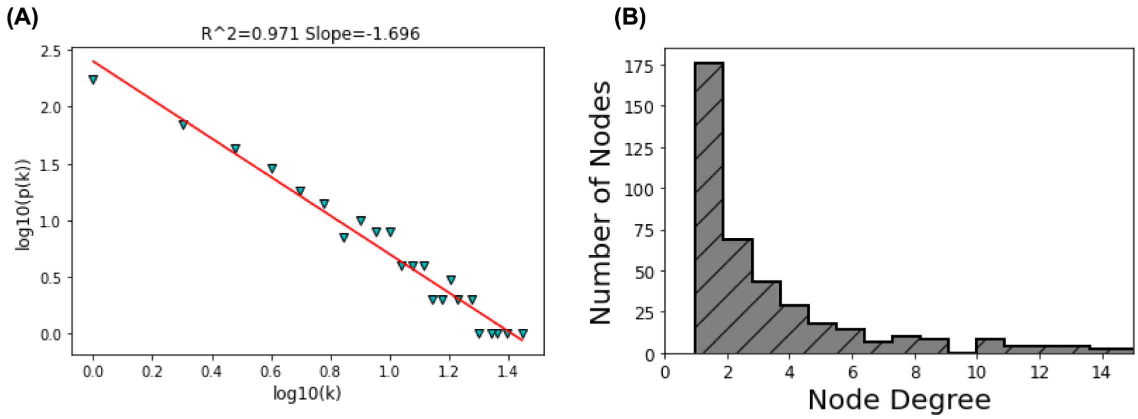

**Figure S1: The static comorbidity network is checked for scale-free topology.** (A) The scale-free topology fitting plot has a coefficient of determination ($R^2$) of 0.97. (B) The distribution of node degrees for the comorbidity network follows a power-law degree distribution.

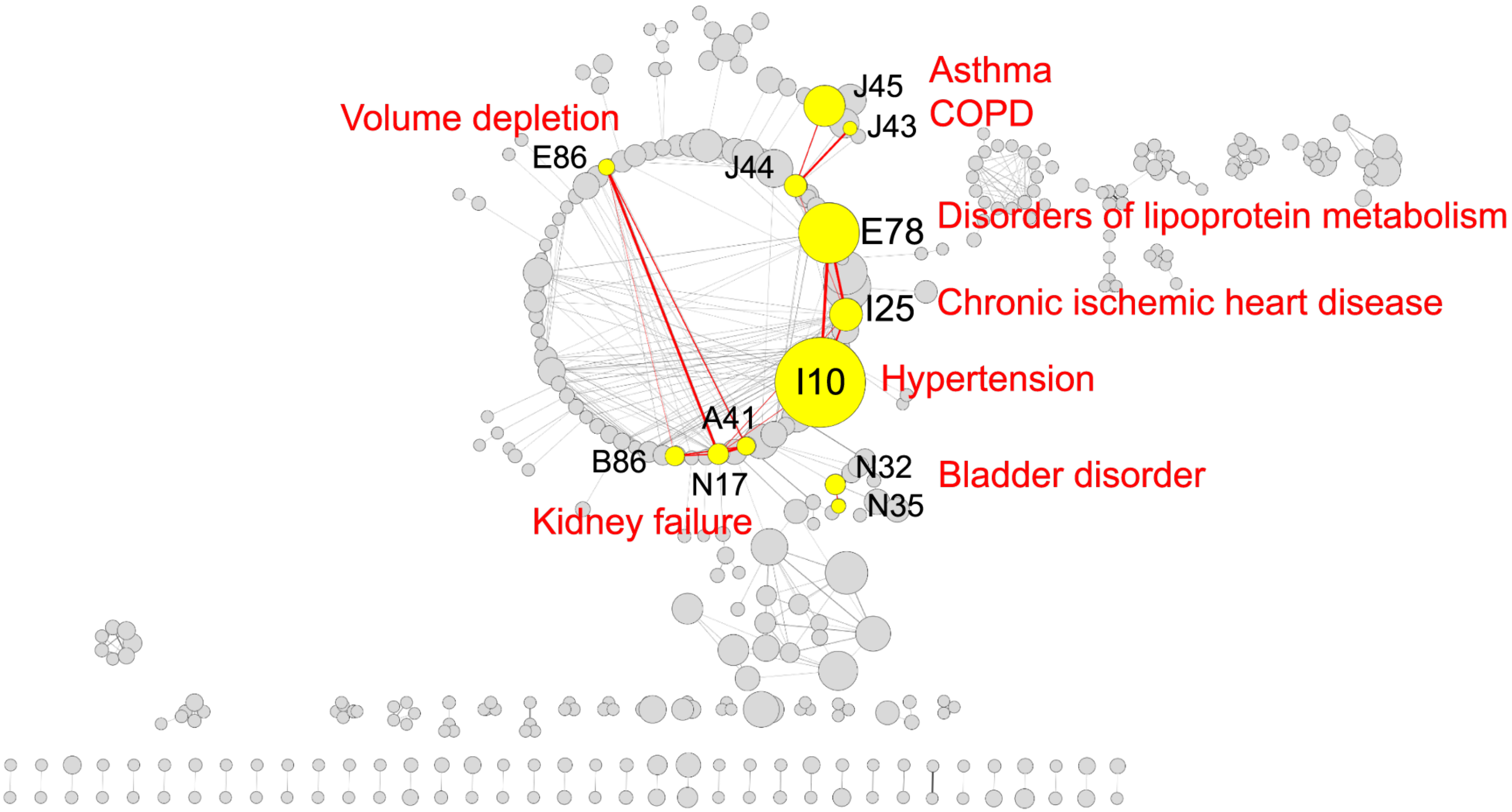

**Figure S2: Comorbidity network with highlighted chronic diseases.** Network visualization of coexisting chronic diseases with common conditions emphasized in red text, showing their co-occurrences in UK Biobank patient histories.

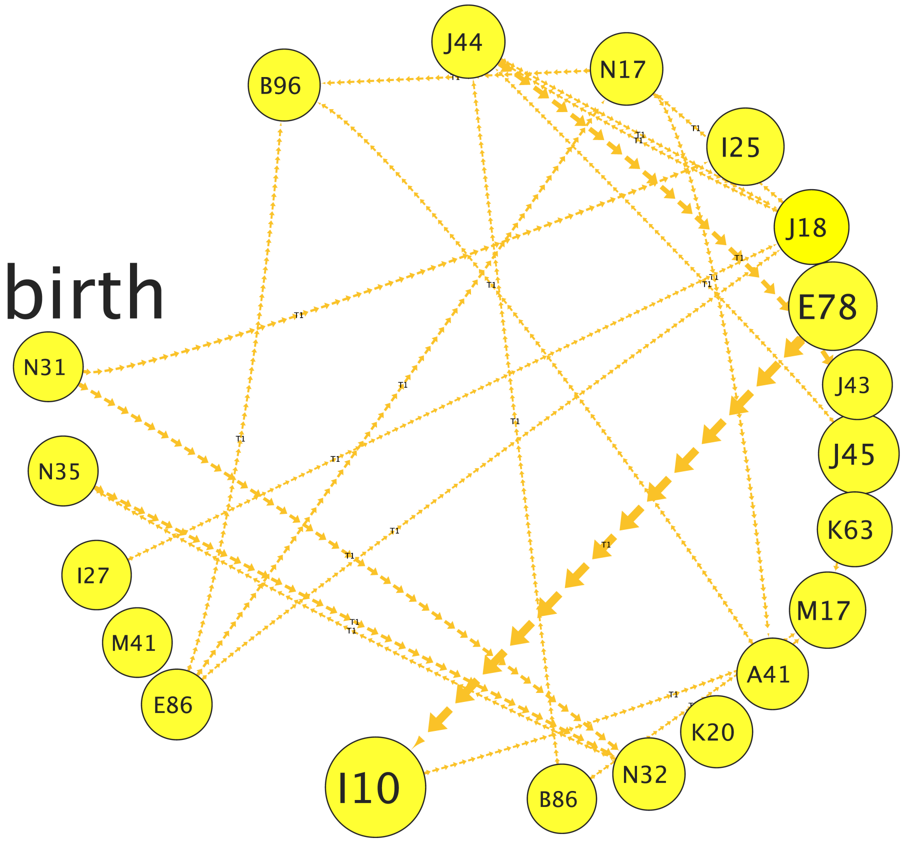

Figure S3: Temporal network illustrating disease co-occurrence within a span of less than 2 weeks.

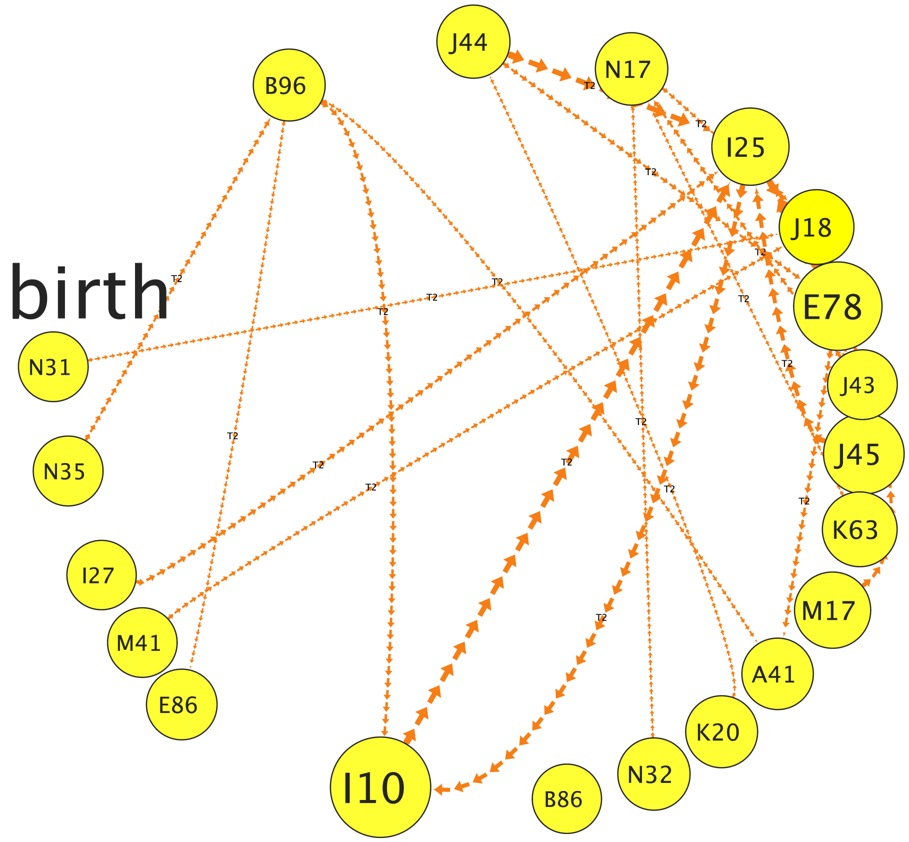

**Figure S4: Directed network depicting disease co-occurrence within a period ranging from 2 weeks to 1 month.**

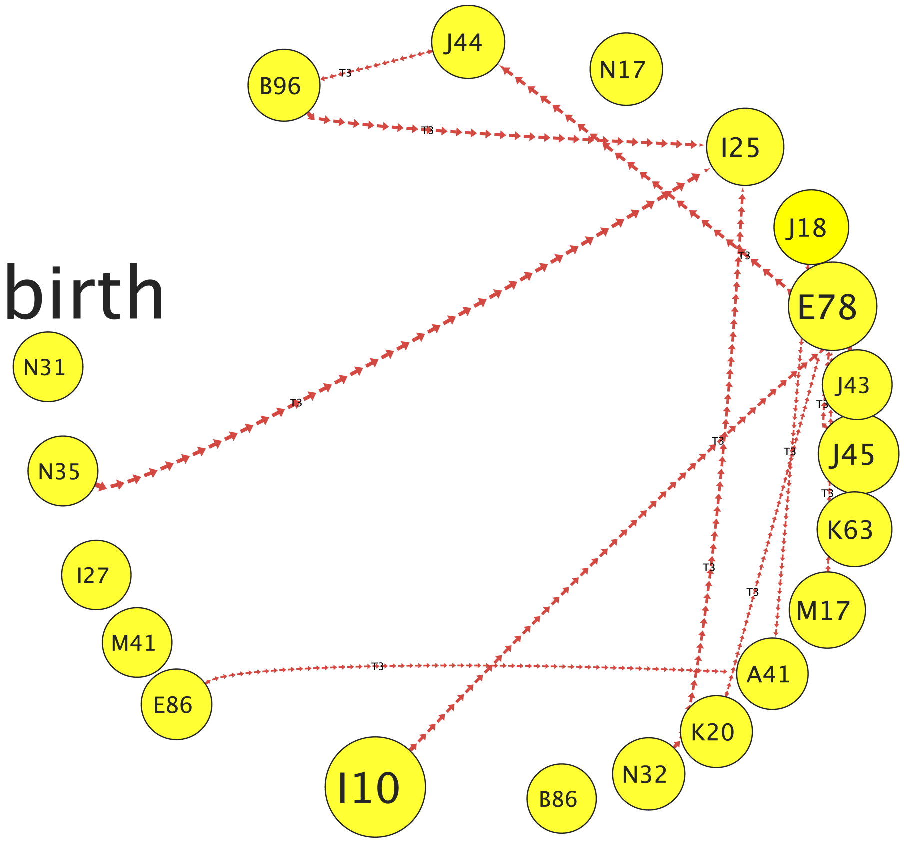

**Figure S5: Temporal disease network illustrating co-occurrence within a 1 to 3 months period.**

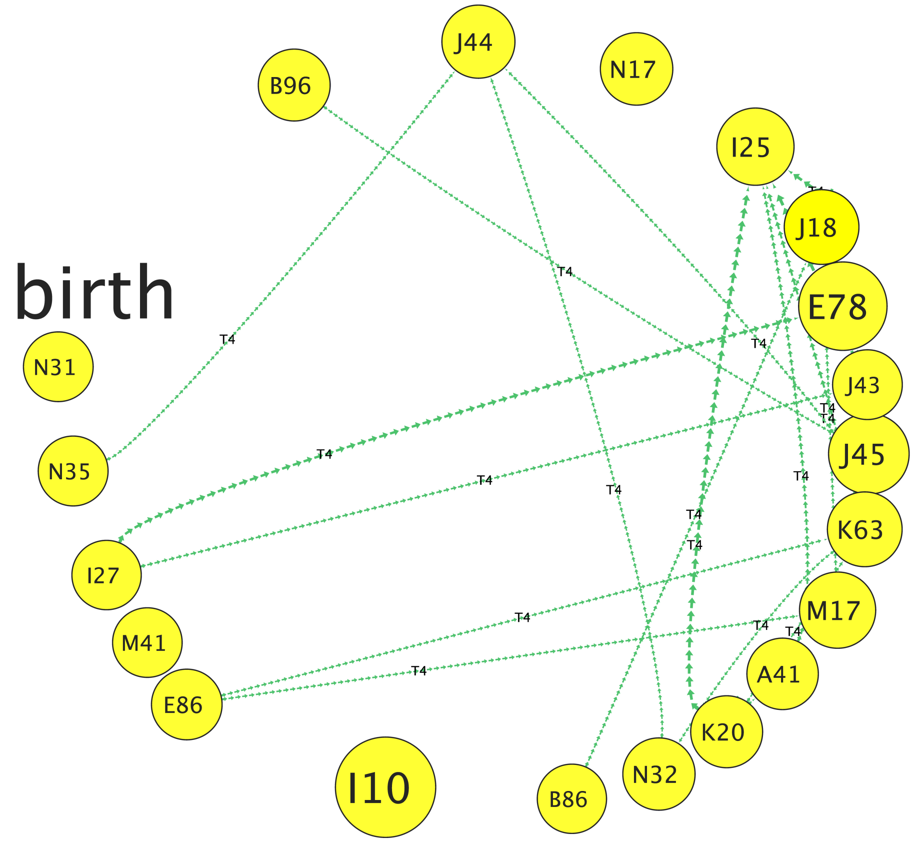

**Figure S6: Temporal disease network illustrating co-occurrence within a 3 to 6 months period.**

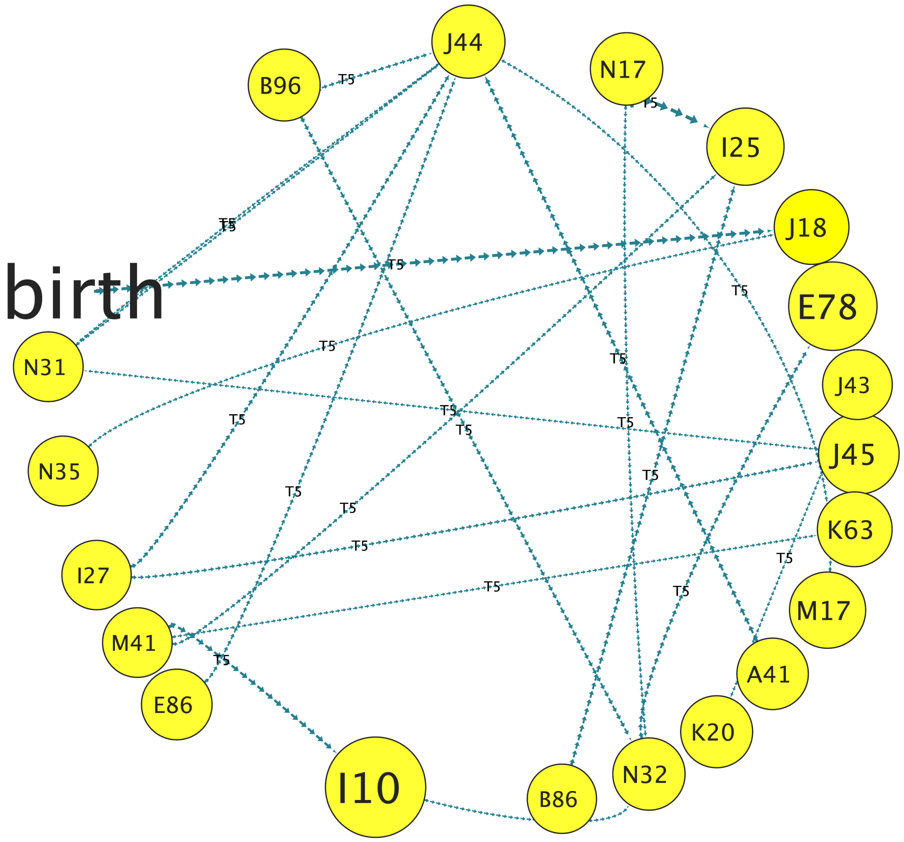

Figure S7: Temporal disease network depicting co-occurrence within a 6-month to 1-year period.

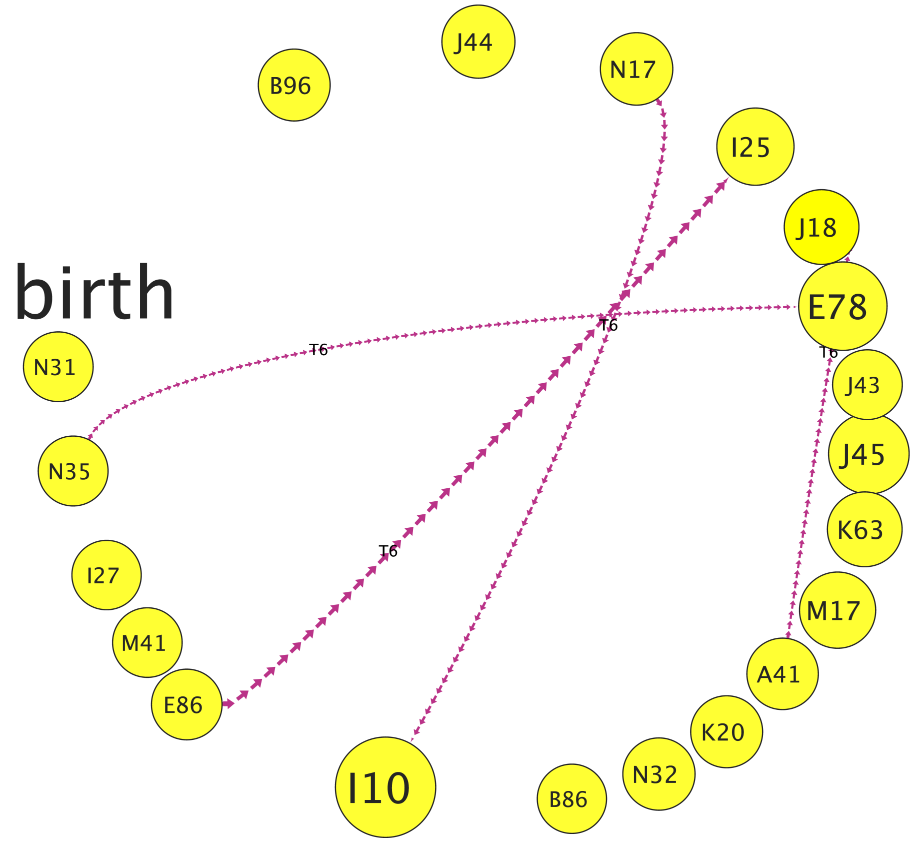

Figure S8: Temporal comorbidity network illustrating disease co-occurrence within 1 to 2 years.

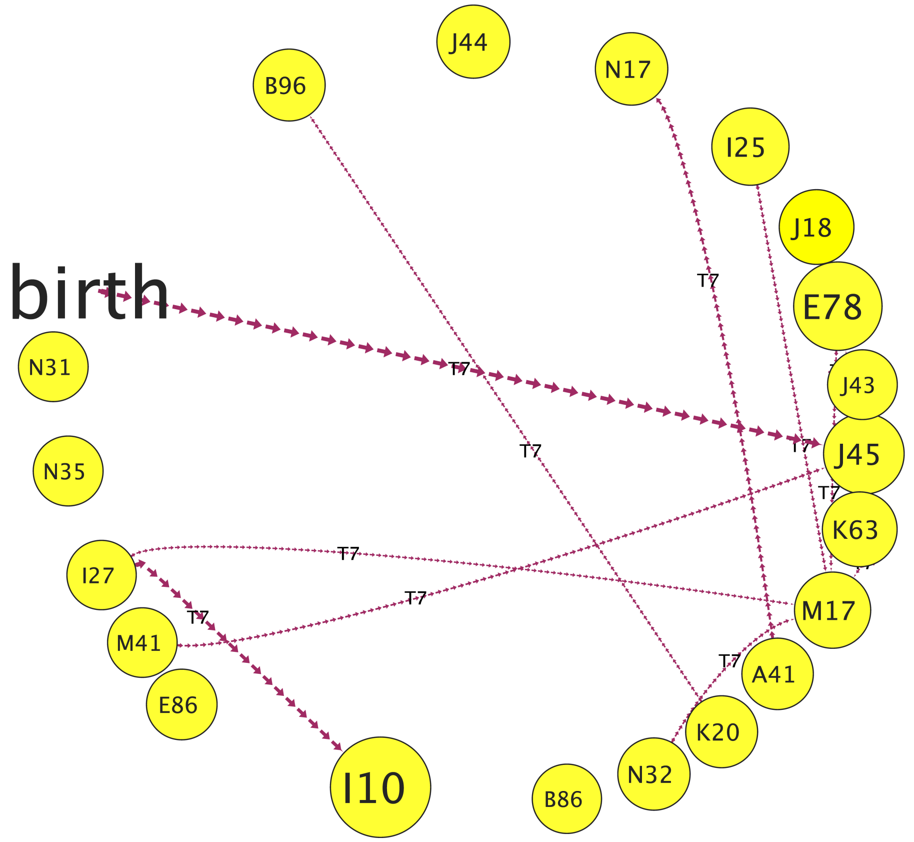

Figure S9: Temporal disease network highlighting disease co-occurrence within 2 to 5 years.

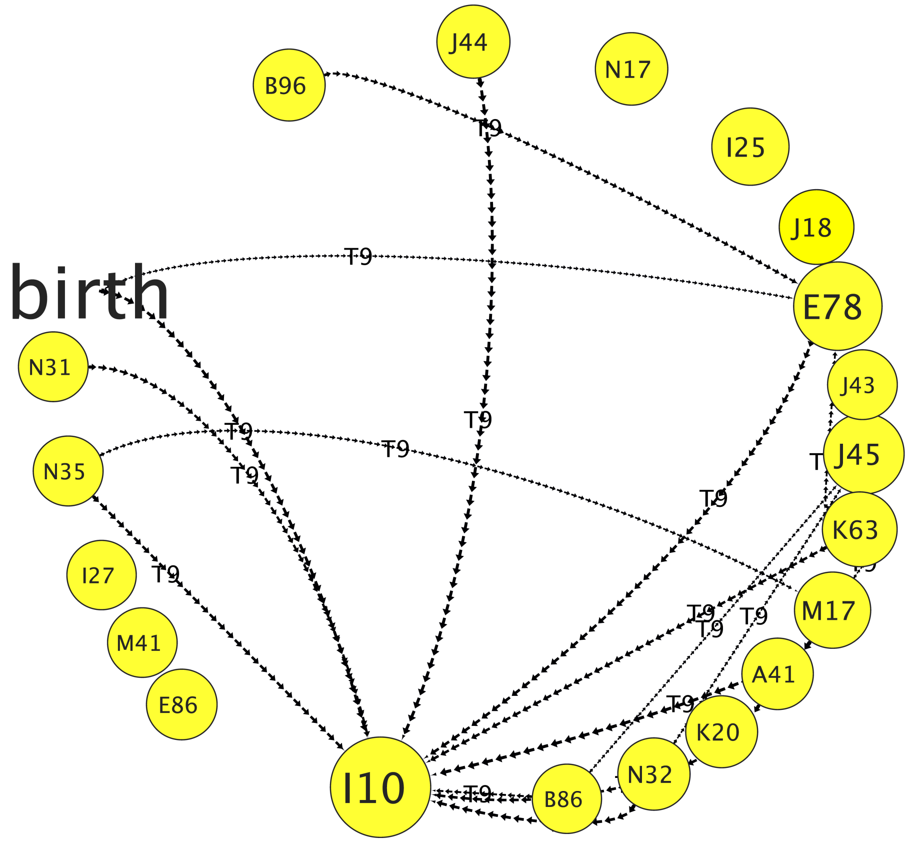

Figure S10: Temporal disease network illustrating disease co-occurrences spanning a period of more than 15 years.
